# Corticostriatal connectivity mediates the reciprocal relationship between sleep and impulsivity in early adolescents

**DOI:** 10.1101/2022.11.07.22282025

**Authors:** Fan Nils Yang, Tina Tong Liu, Ze Wang

## Abstract

**Background:** Adolescence, a developmental period characterized by major changes in sleep and circadian rhythms, is associated with normative increases in impulsivity. While insufficient sleep has been linked to elevated impulsivity, the neural mechanism underlying the relationship remains poorly understood.

**Methods:** We analyzed a dataset of 7,884 drug-naive 9-10 year-olds from the Adolescent Brain Cognitive Development (ABCD) study. Among them, 5,166 have 2-year follow-up neuroimaging data. Linear mixed-effects models, mediation analysis, and longitudinal mediation analysis were used to investigate the relationship between sleep, impulsivity, and brain functional and structural connectivity between the cortex and the striatum.

**Results:** We found that less sleep is significantly associated with higher impulsivity and disrupted functional connectivity between the cingulo-opercular network and the left caudate, and between the cingulo-parietal network and the right pallidum. These two connectivity measurements mediate the effect of sleep duration on impulsivity at both baseline and two-year follow-up. Longitudinal mediation analyses further revealed that sleep duration and impulsivity can reinforce each other through cortical-striatum connectivities in a reciprocal manner.

**Conclusions:** These results reveal neural mechanisms underlying the robust reciprocal relationship between insufficient sleep and impulsivity. Our findings highlight the role of early sleep intervention in helping early adolescents control their impulses, which might in turn prevent the development of substance use.

## Introduction

As part of the normative development, adolescence is a crucial period for brain and affective development, accompanied by increased sensitivity to reward, risk-taking, and impulsivity (Crone and van Duijvenvoorde, 2021; Dahl, 2004; van den Bos et al., 2015). Adequate sleep is essential for brain, cognitive, and affective development during this period (Cheng et al., 2020; Dang-Vu et al., 2006; Kopasz et al., 2010; Maret et al., 2011; Wang et al., 2011; Yang et al., 2022b). Yet, more than 50% of 9-10 year-olds sleep less than the recommended amount (i.e., nine hours of sleep per day) in the United States (Yang et al., 2022b). Previous literature suggests a bidirectional relationship between insufficient sleep and impulsivity in young adolescents (Bauducco et al., 2019). Given the prevalence of insufficient sleep in early adolescents, it is crucial to understand the neural circuitry that mediates the relationship between sleep and impulsivity.

Despite the vital importance of sleep, only a few studies have investigated the effect of insufficient sleep on brain functions or structures in adolescents using magnetic resonance imaging techniques (Cheng et al., 2020; Dutil et al., 2018; Yang et al., 2022b). For example, higher variability in sleep duration was associated with reduced structural connectivity (fractional anisotropy, FA) in the frontostriatal tract one year later (Telzer et al., 2015). Moreover, by using a population-based sample and propensity score matching method, Yang et al. (2022) provide strong evidence that insufficient sleep has long-lasting impacts on cortico-basal ganglia connectivity (Yang et al., 2022b). These studies converged to suggest that the striatum (and basal ganglia more broadly) plays an important role in irregular or insufficient sleep. Indeed, lesions to the striatum in rats led to sleep disturbances, including a marked reduction in slow-wave sleep and sleep duration (Mena-Segovia et al., 2002; Qiu et al., 2010). Striatum is a key component of cortex-basal ganglia loops and is heavily implicated in impulsivity and substance use (Koob and Volkow, 2010). It is then possible that reduced corticostriatal connectivity following insufficient sleep might underlie the problem of impulsive behaviors in early adolescents.

To this end, we investigated how sleep duration impacts corticostriatal connectivity in early adolescents over two years. Specifically, we examined functional and structural connectivity measurements identified in Yang et al. (2022b) and impulsivity measured by the Impulsive Behaviour Scale for children. Using a population-based sample of more than 11,000 9-10 year-olds in the Adolescent Brain Cognitive Development (ABCD) study, we tested the hypothesis that insufficient sleep is associated with (i) reduced functional and structural connectivities between the striatum and cortex and (ii) higher impulsivity. We further predicted that the identified corticostriatal connectivity mediates the effect of sleep duration on impulsivity.

## Methods

### 2.1 Data source

We used ABCD data release 4.0 (released in September 2021), which includes behavioral and neuroimaging data from 11,876 children collected at baseline and 2-year follow-up. Detailed protocols of the ABCD study were previously reported (Casey et al., 2018). The ABCD data were collected from a nationally distributed set of 21 recruitment sites and approved by institutional review boards (IRB) at the University of California, San Diego for the ethical review and approval of the research protocol. Written informed consent from parents and assent from children were obtained at each site. Recruitment reflected the distributions of demographic and socio-economic characteristics in the U.S. population of 9–10 year-old children (sex, race, ethnicity, household income, etc.). Children with serious neurological or psychiatric diagnoses were excluded. Detailed information about the protocols of the ABCD study can be found elsewhere (https://abcdstudy.org/scientists/protocols/) (Casey et al., 2018).

Among all children in the ABCD study, 7,884 passed the following two inclusion criteria in the current study (see **Figure S1**). First, they must pass both resting-state functional MRI and DTI quality control provided by the ABCD study. Second, they had not used any kind of substance or alcohol based on the ABCD Youth Substance Use Interview. Out of these 7,884 children, 5,166 had 2-year follow-up data. see **Table 1** for detailed demographic information.

**Table 1.** Demographic information of the included participants from the ABCD dataset.

| Characteristic | Baseline | Two-year Follow-up |
| --- | --- | --- |
|  | N (%) or Mean (SD) | N (%) or Mean (SD) |
| Age (month) | 119.3 (7.6) | 143.6 (7.9) |
| Sex at birth |  |  |
| female | 3937 (49.9%) | 2483 (48.1%) |
| male | 3947 (50.1%) | 2683 (51.9%) |
| Race |  |  |
| White | 5161 (65.5%) | 3534 (68.4%) |
| Black | 1145 (14.5%) | 652 (12.6%) |
| Asians | 165 (2.1%) | 88 (1.7%) |
| Mixes | 1413 (17.9%) | 892 (17.3) |
| Pubertal Status | 1.8 (0.9)<sup>1</sup> | 2.6 (1.1)<sup>2</sup> |
| Parent substance use | 2751 (34.9%) | 1855 (35.9%) |
| Substance use during pregnancy | 2582 (32.7%) | 1710 (33.1%) |
| Number of children | 7884 | 5166 |

Note:
1. 268 children have missing values; 2. 4444 children have missing values, as puberty status were not collected after pandemic;

### 2.2 Sleep measures

We estimated the independent variable, a child’s sleep duration, based on the parent-reported Sleep Disturbance Scale for Children administered at baseline and 2-year follow-up. Specifically, we used answers to the question, “How many hours of sleep does your child get on most nights in the past six months?”, which provide estimates of a child’s sleep duration in five categories (≥9 h, 8–9 h, 7–8 h, 5–7 h, <5 h). As only a few children (n=22) had less than 5 hours of sleep per day, children with 5-7 h and less than 5 h of sleep were grouped as less than 7 hours of sleep group.

### 2.3 Impulsivity measures

We estimated a child’s five different aspects of impulsivity as dependent variables on the basis of scores from the Urgency, Premeditation (lack of), Perseverance (lack of), Sensation Seeking, and Positive Urgency (UPPS-P) Impulsive Behaviour Scale for children (ABCD version). Specifically, Negative Urgency means “tendency to act rashly under extreme negative emotions”; Positive Urgency: “tendency to act rashly under extreme positive emotions”; Lack of Premeditation: “tendency to act without thinking”; Lack of Perseverance: “inability to remain focused on a task”; Sensation Seeking: “tendency to seek out novel and thrilling experiences”.

### 2.4 Brain measures

All children had a standardized set of a structural MRI scan, four resting-state functional MRI scans, and a diffusion tensor imaging scan at baseline and 2-year follow-up. Acquired images were preprocessed and controlled for quality purposes at the Data Analysis, Informatics, and Resource Centre of the ABCD study (Hagler et al., 2019). Site effects were harmonized by the ComBat method (Johnson et al., 2007; Yang et al., 2021; Yu et al., 2018). Our previous results show that insufficient sleep has a long-lasting impact on resting-state functional connectivity (rs-FC) between the cingulo-opercular network and left caudate (cerc-cdelh) and between cingulo-parietal network and right pallidum (copa_plrh), and FA of two structural connectivity in superior-corticostriate tract (coticostrate) and superior-corticostriate-parietal tract (coticostriate-parietal) (Yang et al., 2022b). Given the involvement of striatum/basal ganglia in these connectivity measurements, we hypothesize these measurements would also be associated with impulsivity. Brain connectivity measurements used in the current study were calculated and provided by the ABCD study. Detailed scan protocols and processing steps can be found elsewhere (Hagler et al., 2019).

### 2.5 Substance use measures

Substance use history, measured by ABCD Youth Substance Use Attitudes Questionnaire, was used as an exclusion criterion to ensure the population studied is drug-naive, as drug experience might alternate the striatum circuit. Family substance use history (ABCD Parent Family History Summary Scores) and substance use during pregnancy (ABCD Developmental History Questionnaire) were used as covariates. For family substance use history, we coded whether biological parents have any kind of substance use and/or alcohol problems (0 equals none of the parents has any problems, 0.5 means one of the parents has at least one kind of substance use problem, and 1 represents both parents have at least one kind of substance use problems). For substance use during pregnancy, 1 means at least one kind of substance and/or alcohol was used during pregnancy, and 0 means no substance was used during pregnancy.

### 2.6 Statistical analyses

Linear Mixed-effects Models (LME, implemented through function fitlme in Matlab) were used to investigate the effect of sleep duration on impulsivity, rs-FC, and FA. All models included fixed-effect covariates for age, sex at birth, race (Black, White, Asian, and Mixes or Others), pubertal status (1–4, assessed by ABCD Youth Pubertal Development Scale and Menstrual Cycle Survey History), average motion during the resting scans (mean frame-wise displacement, FD), number of fMRI time points remained after preprocessing, household income, parents’ educational level, body mass index (BMI), family substance use history, substance use during pregnancy, and random effects for family relatives nested within data collection sites. The false discovery rate (FDR, alpha set to 0.05) was applied to correct for multiple comparisons.

Motivated by the findings that brain measures mediate the relationship between sleep patterns and development outcomes (Yang et al., 2022a), we first tested whether baseline brain measures can mediate the effect of sleep duration on impulsivity. Mediation analyses were performed using an established neuroimaging mediation toolbox (Wager et al., 2009). Statistical significance was assessed by 10,000 random-generated bootstrapped samples. The details of the mediation analysis can be found in Yang et al., 2022a. The aforementioned covariates were controlled in all mediation models.

We then explored the longitudinal relationship between sleep duration, impulsivity, and brain connectivity using longitudinal mediation analyses. Specifically, we tested whether the identified brain connectivity measurements can mediate (i) the relationship between sleep duration at baseline and impulsivity at two-year follow-up (FL2) after controlling for baseline impulsivity and (ii) the association between impulsivity at baseline and sleep duration at FL2 after controlling for baseline sleep duration. The aforementioned covariates were controlled in these mediation models.

## Results

We first examined how sleep duration affects each of the five aspects of impulsivity from the UPPS-P Impulsive Behaviour Scale, including negative urgency, positive urgency, lack of perseverance, lack of premeditation, and sensation seeking (see **Figure 1A**). We found that sleep duration was significantly associated with negative urgency (*t* = -3.02, FDR corrected *p* < 0.01) and positive urgency (*t* = -4.30, FDR corrected *p* < 0.001), but not with lack of premeditation, lack of perseverance, and sensation seeking (all FDR corrected *p* > 0.05).

**Figure 1.**
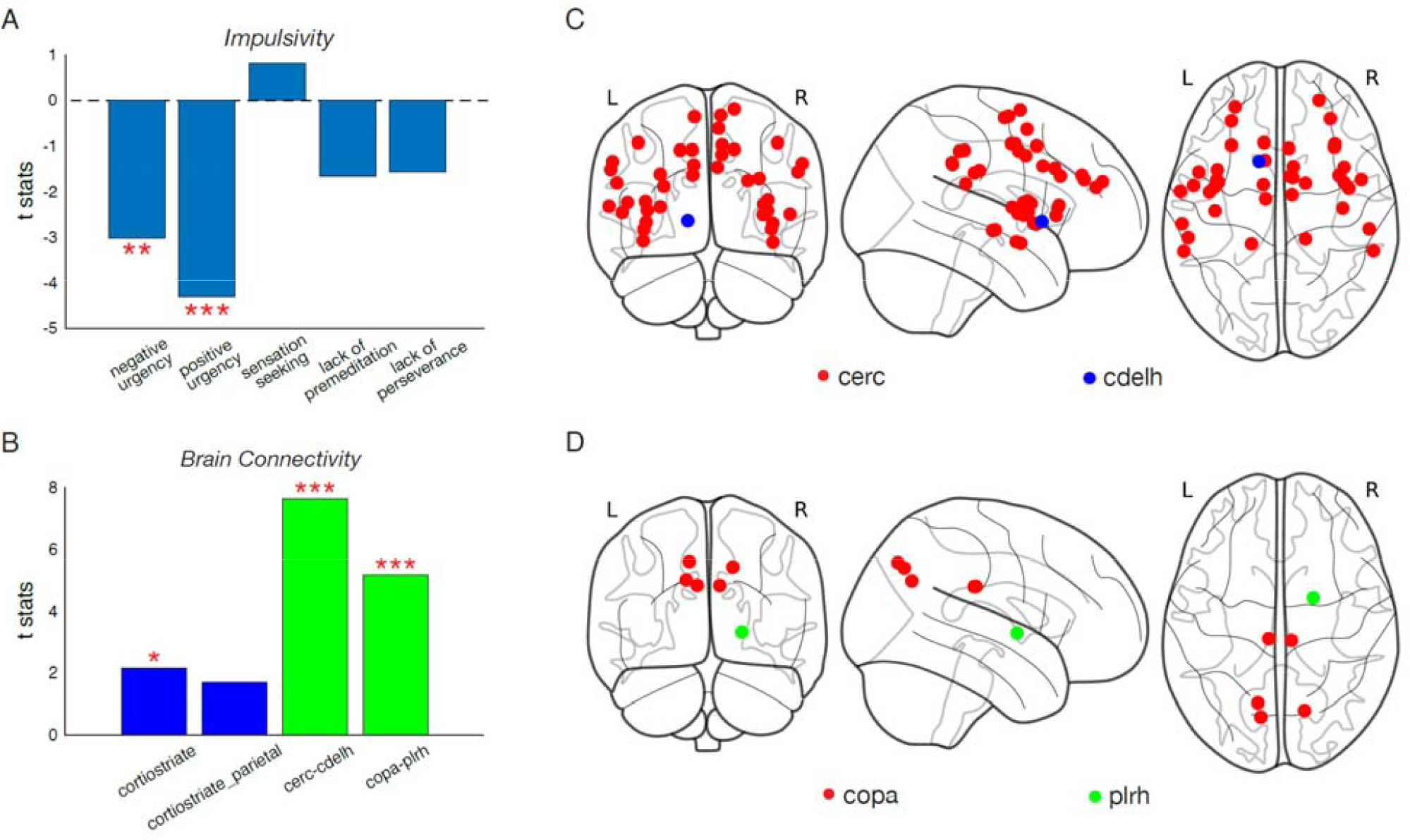
The relationship between sleep duration and impulsivity/brain connectivity. (A) The strength of the relationship between sleep duration and measures of impulsivity. (B) The strength of the relationship between sleep duration and structural connectivity (blue) and between sleep duration and functional connectivity (green). (C) Visualization of nodes in the cingulo-opercular network (red) and left caudate (blue). (D) Visualization of nodes in the cingulo-parietal network (red) and right pallidum (green). Note: * denotes *p* < 0.05; ** denotes *p* < 0.01, *** denotes *p* < 0.001. Cerc: cingulo-opercular network; cdelh: left caudate; copa: cingulo-parietal network; plrh: right pallidum.

Next, we examined how sleep duration affects structural and functional connectivity (**Figure 1B**). Out of the two structural connectivity measures (in blue) and two functional connectivity measures (in green), corticostrate tract (*t* = 2.15, FDR corrected *p* < 0.05), functional connectivity between cingulo-opercular network and the left caudate, (cerc-cdelh, *t* = 7.65, FDR corrected *p* < 0.001), and functional connectivity between cingulo-parietal network and the right pallidum (copa-plrh, *t* = 5.17, FDR corrected *p* < 0.001) were significantly associated with sleep duration. See **Figure 1C & 1D** for the illustration of cerc-cdelh and copa-plrh.

To examine whether the effects of sleep duration on negative urgency and positive urgency were contingent on the identified brain connectivity, we performed mediation analyses (**Figure 2A & 2C**) and found that (i) cerc-cdelh connectivity significantly mediated the effect of sleep duration on negative urgency (beta = -0.014, bootstrapped *p* < 0.05, **Figure 2B**) and positive urgency (beta = -0.027, bootstrapped *p* < 0.001, **Figure 2B**), (ii) copa-plrh significantly mediated the effect of sleep duration on negative urgency (beta = -0.009, bootstrapped *p* < 0.05, **Figure 2D**) and positive urgency (beta = -0.018, bootstrapped *p* < 0.001, **Figure 2D**), and (iii) corticostrate did not mediate the effect of sleep duration on both impulsivity measures (bootstrapped *p* >0.05).

**Figure 2.**
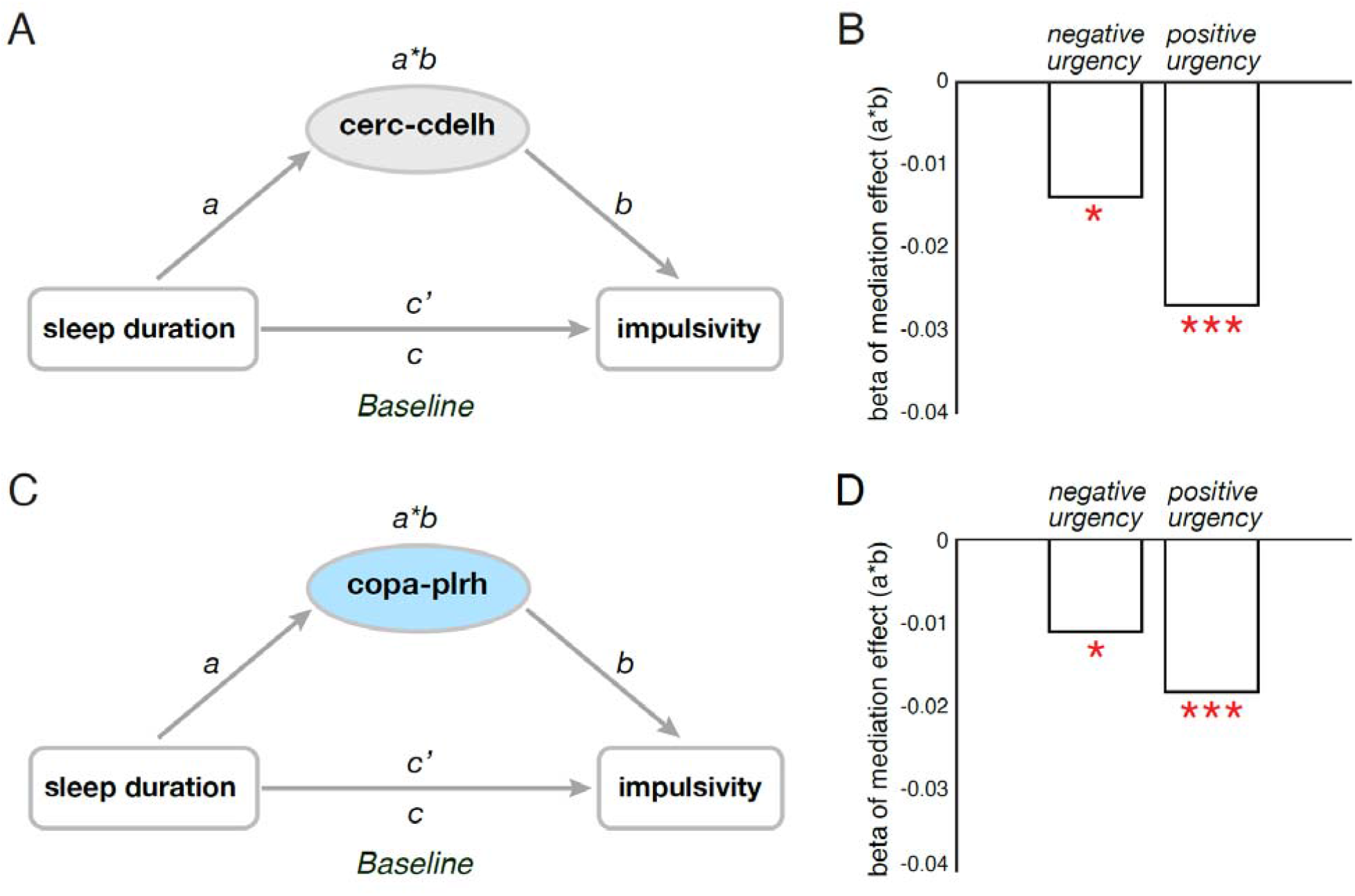
At baseline, cortico-striatal functional connectivity measures mediated the relationship between sleep duration and impulsivity. (A) Diagram of the mediation models. (B) Mediation effects of cerc-cdelh on the associations between sleep duration and positive and negative urgency. (C) Diagram of the mediation models. (D) Mediation effects of copa-prlh on the associations between sleep duration and ugency. Note: * denotes *p* < 0.05; *** denotes *p* < 0.001; cerc-cdelh: functional connectivity between cingulo-opercular network and left caudate; copa-plrh: functional connectivity between cingulo-parietal network and right pallidum.

Similarly, we found that insufficient sleep at FL2 is significantly associated with elevated positive and negative urgency at FL2 and reduced cerc-cdelh and copa-plrh at FL2 (See **Figure S2**). For mediation analysis, both cerc_cdelh and copa_plrh at FL2 mediate the effect of sleep duration at FL2 on positive urgency at FL2 but not on negative urgency at FL2 (see **Figure 3**). In addition, paired t-test revealed that three of the five impulsivity measures, including positive and negative urgency and sensation seeking, decreased over the two years (see **Figure S3**).

**Figure 3.**
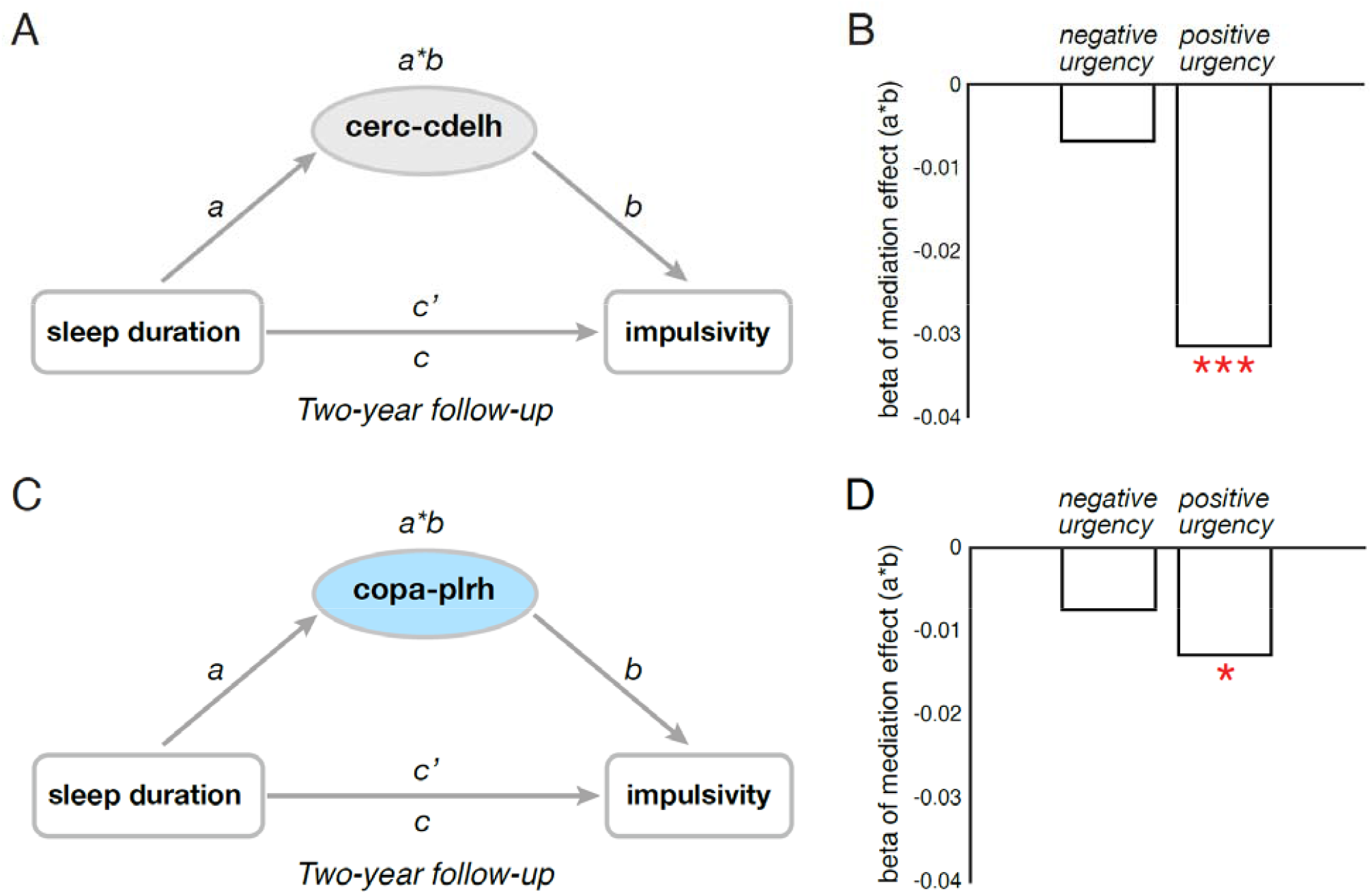
At two-year follow-up, cortico-striatal functional connectivity measures mediated the relationship between sleep duration and impulsivity. (A) Diagram of the mediation models. (B) Mediation effects of cerc-cdelh on the associations between sleep duration and positive and negative urgency. (C) Diagram of the mediation models. (D) Mediation effects of copa-prlh on the associations between sleep duration and ugencies. Note: * denotes *p* < 0.05; *** denotes *p* < 0.001; cerc-cdelh: functional connectivity between cingulo-opercular network and left caudate; copa-plrh: functional connectivity between cingulo-parietal network and right pallidum.

Finally, longitudinal mediation analysis revealed that both rs-FC measurements (cerc_cdelh and copa_plrh) mediated the effect of sleep duration at baseline on positive urgency at FL2, after controlling for baseline positive urgency (bootstrapped *p* < 0.05, see **Figure 4A&4B**). Similarly, both connectivity also mediated the effect of positive urgency at baseline on sleep duration at FL2, after controlling for baseline sleep duration (bootstrapped *p* < 0.05, see **Figure 4C&4D**).

**Figure 4.**
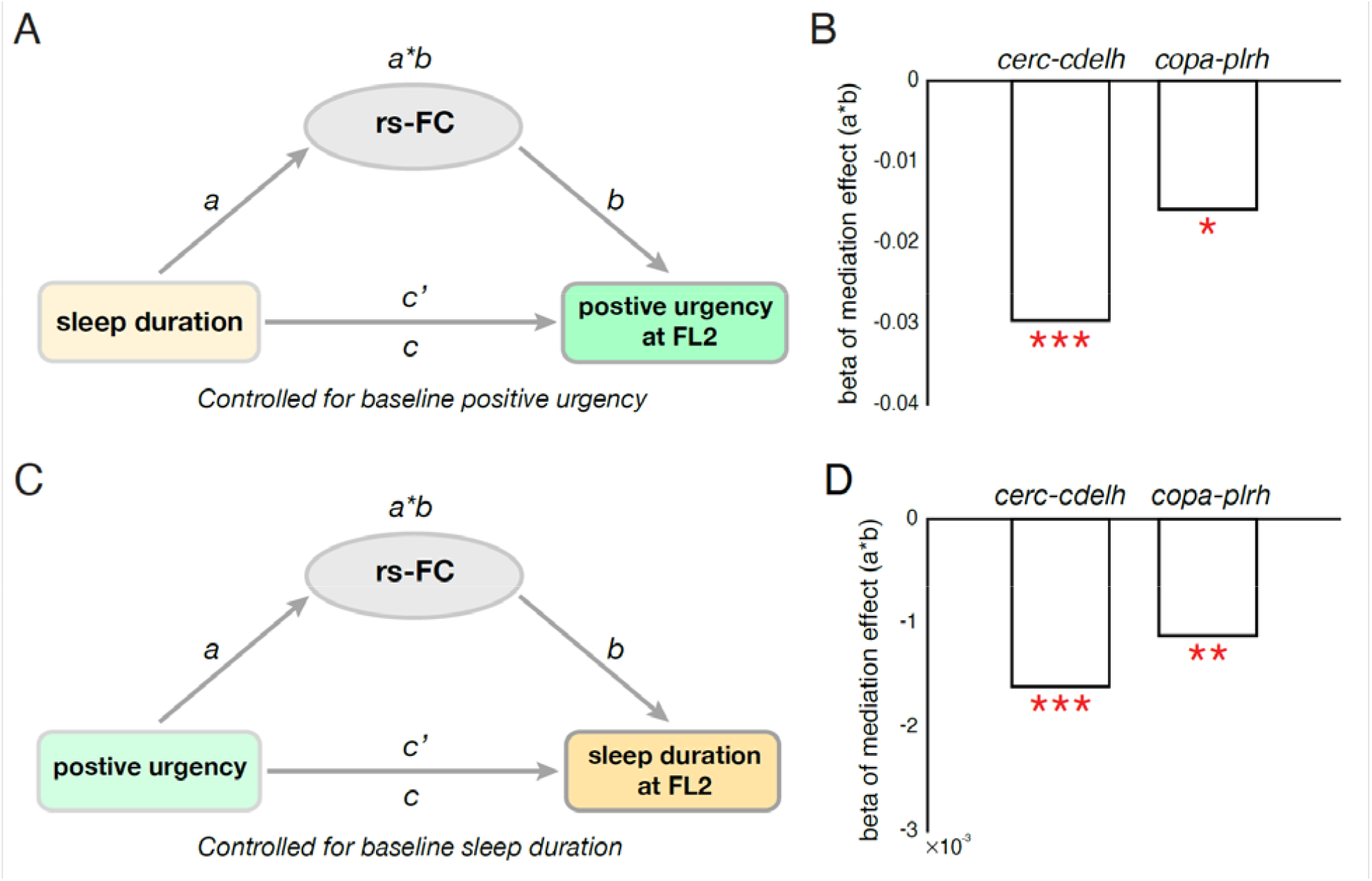
Longitudinal mediation analysis revealed cortico-striatal functional connectivity measures mediated the reciprocal relationship between sleep duration and impulsivity. (A) Diagram of the longitudinal mediation models: the independent variable is sleep duration at baseline, mediators are two functional connectivity measurements, the dependent variable is positive urgency at FL2. (B) Mediation effects of cerc-cdelh or copa-plrh on the associations between sleep duration and positive urgency at FL2. (C) Diagram of the longitudinal mediation models: the independent variable is positive urgency at baseline, mediators are two functional connectivity measurements, the dependent variable is sleep duration at FL2. (D) Mediation effects of cerc-cdelh or copa-plrh on the associations between positive urgency and sleep duration at FL2. Note: * denotes *p* < 0.05; ** denotes *p* < 0.01, *** denotes *p* < 0.001; cerc-cdelh: functional connectivity between cingulo-opercular network and left caudate; copa-plrh: functional connectivity between cingulo-parietal network and right pallidum.

## Discussion

By controlling for various key covariates related to impulsivity and sleep, we demonstrated a potential neural mechanism underlying the effect of sleep duration on impulsivity in drug-naive children. First, we found that two cortico-striatal connectivity measurements mediate the effect of sleep duration on the effective aspect of impulsivity, i.e., negative and positive urgency. Second, the longitudinal analysis revealed that sleep duration and positive urgency at baseline can predict each other two years later through these two cortico-striatal connectivity measurements. Our findings shed light on using early sleep intervention to control impulses in early adolescents.

The current results strongly suggest that cortico-striatal connectivity, especially the connectivity between the cingulo-opercular network and left caudate, is a neural circuitry underlying the relationship between insufficient sleep and elevated level of impulsivity, specifically negative and positive urgency, in early adolescents. Impulsivity is a multifaceted construct, consisting of separable constructs of affect-related impulsivity, such as urgency and sensation seeking, and cognitive impulsivity, such as lack of premeditation and lack of perseverance (Cyders and Smith, 2008; Whiteside and Lynam, 2001). Given that caudate is involved in attention, cognitive control, reward, and emotion processing (Monk, 2008; Robinson et al., 2012), reduced between caudate and cortical regions after insufficient sleep may underlie increased salience of reward cues and a lack of inhibitory control on emotion, giving rise to excessive urgency. Indeed, a previous study found that negative urgency is related to insufficient involvement of the striatum and anterior cingulate cortex, which are regions that respond to cognitive control and reinforcement learning behavior (Chester et al., 2016). The relationship between negative and positive urgency and insufficient sleep we find is in line with previous findings that affect-related impulsivity is most affected by sleep loss (Kahn-Greene et al., 2007; Kamphuis et al., 2012; Peterson and Benca, 2006)(Peterson and Benca, 2006).

Our longitudinal mediation analysis confirmed the reciprocal relationship between insufficient sleep and urgency over two years in early adolescents. Previous literature suggests that low levels of serotonin (5-HT) and its associated high level of dopamine in the neural system may contribute to the development of urgency (Smith and Cyders, 2016). The potential mechanism undying this reciprocal relationship might be the monoamine neurotransmitters in the striatum circuit. For example, insufficient sleep is associated with decreased sensitivity in 5-HT_1A_ receptors in the brain (Longordo et al., 2009), which plays a role in emotional dysregulation (Gross et al., 2002; Zhuang et al., 1999), and decreased expression of D2/D3 dopaminergic receptors in the striatum, which can subsequently lead to elevated dopamine levels (Longordo et al., 2009). Future work will need to test the neurotransmitter mechanisms, specifically in the basal ganglia/striatum, between insufficient sleep and urgency.

Although adolescence is a period with a normative increase in impulsivity, especially in sensation seeking (Harden and Tucker-Drob, 2011), in the current sample of 9-10-year-olds the levels of impulsivity (i.e., positive urgency, negative urgency, and sensation seeking) decreased over two years. One possible reason is that the basal ganglia system is still immature at the age of 9-12. The basal ganglia are one of the regions that undergo the greatest structural changes in early and late adolescence (Sowell et al., 1999).

Coinciding with basal ganglia maturation, substance use initiation often happens in mid-late adolescence (Johnston et al., 2021). Neurodevelopment models suggest that adolescents’ vulnerability to problematic substance use is attributed to a predominance of a striatum-mediated reward system over a cortically-mediated inhibition control system (Casey et al., 2008; Luna et al., 2015; Steinberg, 2010). However, a recent meta-analysis revealed that substance use risk, e.g. having a family history of substance use, was mostly associated with striatal activation rather than prefrontal/cortical activation during tasks (Tervo-Clemmens et al., 2020). The current study contributes to this literature by controlling the family history of substance use and substance use during pregnancy and demonstrating reduced resting-state cortico-striatal connectivity is associated with elevated impulsivity in drug-naive children. Future studies may test whether cortico-striatal connectivity can predict future substance use onset in adolescents.

A few limitations in the current work should be noted. First, in the current study, we used parent-reported sleep duration, which might not be as accurate as objective sleep duration measured by wearable devices. The ABCD study has begun to collect sleep duration using the Fitbit watch started from two-year follow-up. Future studies might utilize objective sleep duration when more data are available from the ABCD study. Second, we only test functional connectivity between a certain network, i.e. cingulo-opercular network, and striatum. Future studies might test whether specific region-to-region connectivity measurements can mediate the relationship between sleep and impulsivity. Third, other sleep measures, such as sleep quality, might also contribute to abnormal impulsivity during adolescence. For example, Tashjian, et al., (2017) found that poor sleep quality is related to elevated urgency scores and functional connectivity between the prefrontal cortex and default mode network in mid-late adolescence (Tashjian et al., 2017).

Collectively, this study established that cortico-striatal connectivity might be a potential neural mechanism underlying the reciprocal relationship between sleep duration and impulsivity. Given that increased urgency can predict future substance use (Smith and Cyders, 2016), early sleep intervention might be a useful and effective approach to prevent or reduce the risks of substance use in early adolescents.

## Data Availability

All data produced in the present study are available upon reasonable request to the authors

## Data Availability

Scientists can obtain access to individual-level data from the UK Biobank by applying to UK Biobank (https://www.ukbiobank.ac.uk/enable-your-research).

---

**Figure S1.**
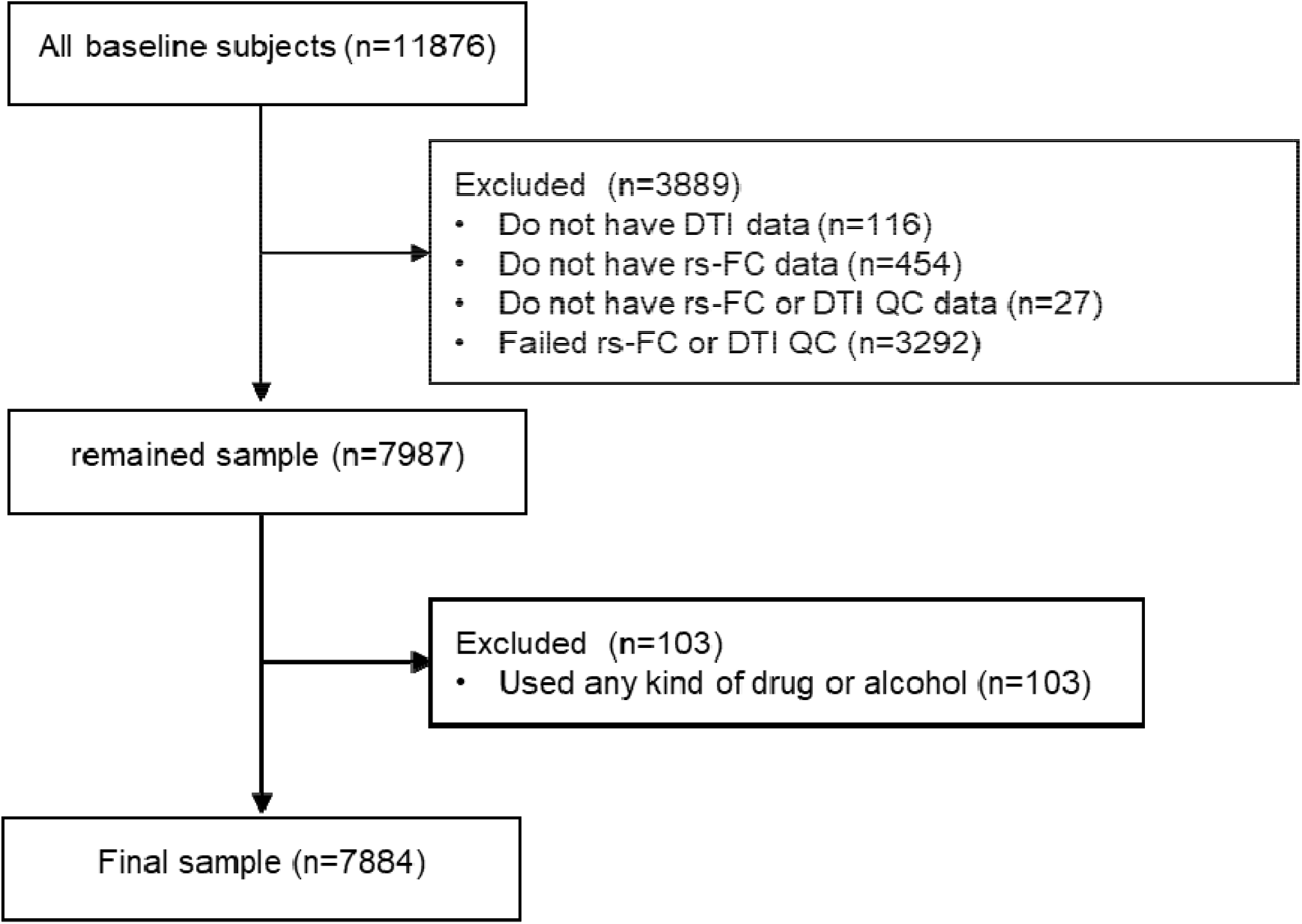
CONSORT diagram about the included and excluded participants.

**Figure S2.** The relationship between sleep duration and impulsivity/brain connectivity at two-year follow-up. Note: ** denotes *p* < 0.01; *** denotes *p* < 0.001; FL2: two-year follow-up.

**Figure S3.**
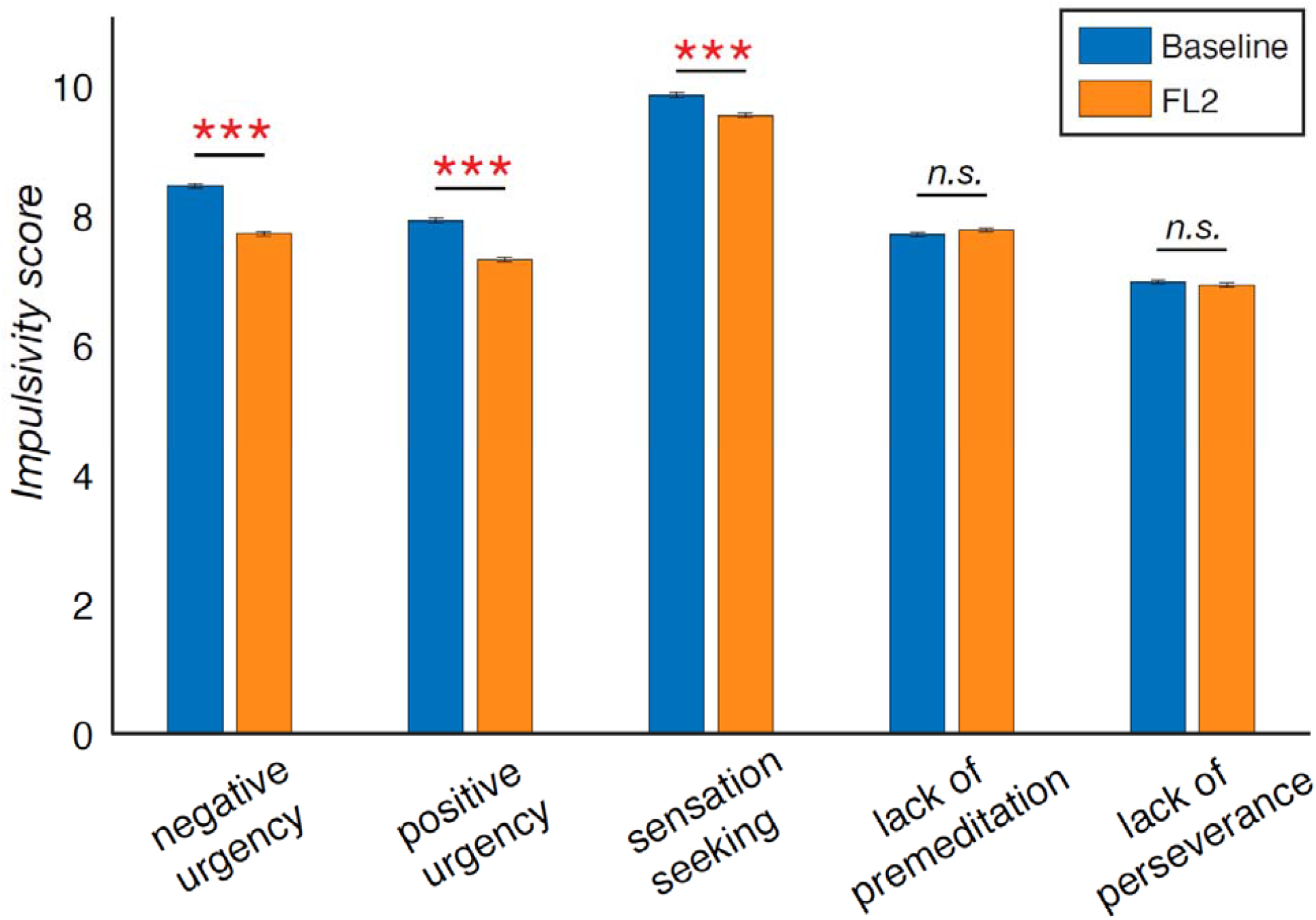
Pair-sample t-tests on how impulsivity changes between baseline and two-year follow-up. Note: ***, *p* < 0.001, n.s., not significant; FL2, two-year follow-up. Error bar: standard error.

## Acknowledgment

Research efforts (data analysis and interpretation) in this work were supported by NIH grants: R01AG060054, R01AG070227, R01EB031080-01A1, P41EB029460-01A1, 1UL1TR003098. We thank the ABCD consortium and NIH for providing the data for performing the research in this work. Data used in the preparation of this article were obtained from the ABCD Study (https://abcdstudy.org/) and are held in the NIMH Data Archive. This is a multisite, longitudinal study designed to recruit more than 10,000 children aged 9–10 and follow them over 10 years into early adulthood. The ABCD Study is supported by the National Institutes of Health (NIH) and additional federal partners under award numbers U01DA041022, U01DA041028, U01DA041048, U01DA041089, U01DA041106, U01DA041117, U01DA041120, U01DA041134, U01DA041148, U01DA041156, U01DA041174, U24DA041123, and U24DA041147. A full list of supporters is available at https://abcdstudy.org/federal-partners/. A listing of participating sites and a complete listing of the study investigators can be found at https://abcdstudy.org/principal-investigators/. ABCD consortium investigators designed and implemented the study and/or provided data but did not necessarily participate in the analysis or writing of this report. This manuscript reflects the views of the authors and may not reflect the opinions or views of the NIH or ABCD consortium investigators. we are not paid to write this article by a pharmaceutical company or other agency.

## Author contributions

F.N.Y. conceptualized the study, analyzed the data, generated figures, and wrote the original draft. T.T.L contributed to conceptualization and figure visualization, and edited the manuscript. Z.W. supervised the study and edited the manuscript.

## Ethics approval statement

ABCD study received ethical approval in accordance with the ethical standards of the 1964 Declaration of Helsinki.

## References

Bauducco, S.V., Salihovic, S., Boersma, K., 2019. Bidirectional associations between adolescents’ sleep problems and impulsive behavior over time. Sleep Med. X 1, 100009. https://doi.org/10.1016/j.sleepx.2019.100009

Casey, B.J., Cannonier, T., Conley, M.I., Cohen, A.O., Barch, D.M., Heitzeg, M.M., Soules, M.E., Teslovich, T., Dellarco, D.V., Garavan, H., Orr, C.A., Wager, T.D., Banich, M.T., Speer, N.K., Sutherland, M.T., Riedel, M.C., Dick, A.S., Bjork, J.M., Thomas, K.M., Chaarani, B., Mejia, M.H., Hagler, D.J., Daniela Cornejo, M., Sicat, C.S., Harms, M.P., Dosenbach, N.U.F., Rosenberg, M., Earl, E., Bartsch, H., Watts, R., Polimeni, J.R., Kuperman, J.M., Fair, D.A., Dale, A.M., ABCD Imaging Acquisition Workgroup, 2018. The Adolescent Brain Cognitive Development (ABCD) study: Imaging acquisition across 21 sites. Dev. Cogn. Neurosci. 32, 43–54. https://doi.org/10.1016/j.dcn.2018.03.001

Casey, B.J., Getz, S., Galvan, A., 2008. The adolescent brain. Dev. Rev., Current Directions in Risk and Decision Making 28, 62–77. https://doi.org/10.1016/j.dr.2007.08.003

Cheng, W., Rolls, E., Gong, W., Du, J., Zhang, J., Zhang, X.-Y., Li, F., Feng, J., 2020. Sleep duration, brain structure, and psychiatric and cognitive problems in children. Mol. Psychiatry 1–12. https://doi.org/10.1038/s41380-020-0663-2

Chester, D.S., Lynam, D.R., Milich, R., Powell, D.K., Andersen, A.H., DeWall, C.N., 2016. How do negative emotions impair self-control? A neural model of negative urgency. NeuroImage 132, 43–50. https://doi.org/10.1016/j.neuroimage.2016.02.024

Crone, E.A., van Duijvenvoorde, A.C.K., 2021. Multiple pathways of risk taking in adolescence. Dev. Rev. 62, 100996. https://doi.org/10.1016/j.dr.2021.100996

Cyders, M.A., Smith, G.T., 2008. Emotion-based dispositions to rash action: Positive and negative urgency. Psychol. Bull. 134, 807–828. https://doi.org/10.1037/a0013341

Dahl, R.E., 2004. Adolescent brain development: a period of vulnerabilities and opportunities. Keynote address. Ann. N. Y. Acad. Sci. 1021, 1–22.

Dang-Vu, T.T., Desseilles, M., Peigneux, P., Maquet, P., 2006. A role for sleep in brain plasticity. Pediatr. Rehabil. 9, 98–118.

Dutil, C., Walsh, J.J., Featherstone, R.B., Gunnell, K.E., Tremblay, M.S., Gruber, R., Weiss, S.K., Cote, K.A., Sampson, M., Chaput, J.-P., 2018. Influence of sleep on developing brain functions and structures in children and adolescents: A systematic review. Sleep Med. Rev. 42, 184–201. https://doi.org/10.1016/j.smrv.2018.08.003

Gross, C., Zhuang, X., Stark, K., Ramboz, S., Oosting, R., Kirby, L., Santarelli, L., Beck, S., Hen, R., 2002. Serotonin1A receptor acts during development to establish normal anxiety-like behaviour in the adult. Nature 416, 396–400. https://doi.org/10.1038/416396a

Hagler, D.J., Hatton, SeanN., Cornejo, M.D., Makowski, C., Fair, D.A., Dick, A.S., Sutherland, M.T., Casey, B.J., Barch, D.M., Harms, M.P., Watts, R., Bjork, J.M., Garavan, H.P., Hilmer, L., Pung, C.J., Sicat, C.S., Kuperman, J., Bartsch, H., Xue, F., Heitzeg, M.M., Laird, A.R., Trinh, T.T., Gonzalez, R., Tapert, S.F., Riedel, M.C., Squeglia, L.M., Hyde, L.W., Rosenberg, M.D., Earl, E.A., Howlett, K.D., Baker, F.C., Soules, M., Diaz, J., de Leon, O.R., Thompson, W.K., Neale, M.C., Herting, M., Sowell, E.R., Alvarez, R.P., Hawes, S.W., Sanchez, M., Bodurka, J., Breslin, F.J., Morris, A.S., Paulus, M.P., Simmons, W.K., Polimeni, J.R., van der Kouwe, A., Nencka, A.S., Gray, K.M., Pierpaoli, C., Matochik, J.A., Noronha, A., Aklin, W.M., Conway, K., Glantz, M., Hoffman, E., Little, R., Lopez, M., Pariyadath, V., Weiss, S.RB., Wolff-Hughes, D.L., DelCarmen-Wiggins, R., Feldstein Ewing, S.W., Miranda-Dominguez, O., Nagel, B.J., Perrone, A.J., Sturgeon, D.T., Goldstone, A., Pfefferbaum, A., Pohl, K.M., Prouty, D., Uban, K., Bookheimer, S.Y., Dapretto, M., Galvan, A., Bagot, K., Giedd, J., Infante, M.A., Jacobus, J., Patrick, K., Shilling, P.D., Desikan, R., Li, Y., Sugrue, L., Banich, M.T., Friedman, N., Hewitt, J.K., Hopfer, C., Sakai, J., Tanabe, J., Cottler, L.B., Nixon, S.J., Chang, L., Cloak, C., Ernst, T., Reeves, G., Kennedy, D.N., Heeringa, S., Peltier, S., Schulenberg, J., Sripada, C., Zucker, R.A., Iacono, W.G., Luciana, M., Calabro, F.J., Clark, D.B., Lewis, D.A., Luna, B., Schirda, C., Brima, T., Foxe, J.J., Freedman, E.G., Mruzek, D.W., Mason, M.J., Huber, R., McGlade, E., Prescot, A., Renshaw, P.F., Yurgelun-Todd, D.A., Allgaier, N.A., Dumas, J.A., Ivanova, M., Potter, A., Florsheim, P., Larson, C., Lisdahl, K., Charness, M.E., Fuemmeler, B., Hettema, J.M., Maes, H.H., Steinberg, J., Anokhin, A.P., Glaser, P., Heath, A.C., Madden, P.A., Baskin-Sommers, A., Constable, R.T., Grant, S.J., Dowling, G.J., Brown, S.A., Jernigan, T.L., Dale, A.M., 2019. Image processing and analysis methods for the Adolescent Brain Cognitive Development Study. NeuroImage 202, 116091. https://doi.org/10.1016/j.neuroimage.2019.116091

Harden, K., Tucker-Drob, E., 2011. Individual Differences in the Development of Sensation Seeking and Impulsivity During Adolescence: Further Evidence for a Dual Systems Model. Dev. Psychol. 47, 739–46. https://doi.org/10.1037/a0023279

Johnson, W.E., Li, C., Rabinovic, A., 2007. Adjusting batch effects in microarray expression data using empirical Bayes methods. Biostatistics 8, 118–127. https://doi.org/10.1093/biostatistics/kxj037

Johnston, L.D., Miech, R.A., O’Malley, P.M., Bachman, J.G., Schulenberg, J.E., Patrick, M.E., 2021. Monitoring the Future National Survey Results on Drug Use, 1975-2020: Overview, Key Findings on Adolescent Drug Use, Institute for Social Research. Institute for Social Research.

Kahn-Greene, E.T., Killgore, D.B., Kamimori, G.H., Balkin, T.J., Killgore, W.D.S., 2007. The effects of sleep deprivation on symptoms of psychopathology in healthy adults. Sleep Med. 8, 215–221. https://doi.org/10.1016/j.sleep.2006.08.007

Kamphuis, J., Meerlo, P., Koolhaas, J.M., Lancel, M., 2012. Poor sleep as a potential causal factor in aggression and violence. Sleep Med. 13, 327–334. https://doi.org/10.1016/j.sleep.2011.12.006

Koob, G.F., Volkow, N.D., 2010. Neurocircuitry of Addiction. Neuropsychopharmacology 35, 217–238. https://doi.org/10.1038/npp.2009.110

Kopasz, M., Loessl, B., Hornyak, M., Riemann, D., Nissen, C., Piosczyk, H., Voderholzer, U., 2010. Sleep and memory in healthy children and adolescents–a critical review. Sleep Med. Rev. 14, 167–177.

Longordo, F., Kopp, C., Lüthi, A., 2009. Consequences of sleep deprivation on neurotransmitter receptor expression and function. Eur. J. Neurosci. 29, 1810–1819.

Luna, B., Marek, S., Larsen, B., Tervo-Clemmens, B., Chahal, R., 2015. An Integrative Model of the Maturation of Cognitive Control. Annu. Rev. Neurosci. 38, 151–170. https://doi.org/10.1146/annurev-neuro-071714-034054

Maret, S., Faraguna, U., Nelson, A.B., Cirelli, C., Tononi, G., 2011. Sleep and wake modulate spine turnover in the adolescent mouse cortex. Nat. Neurosci. 14, 1418–1420. https://doi.org/10.1038/nn.2934

Mena-Segovia, J., Cintra, L., Prospéro-Garcı, O., Giordano, M., 2002. Changes in sleep– waking cycle after striatal excitotoxic lesions. Behav. Brain Res. 136, 475–481. https://doi.org/10.1016/S0166-4328(02)00201-2

Monk, C.S., 2008. The development of emotion-related neural circuitry in health and psychopathology. Dev. Psychopathol. 20, 1231–1250. https://doi.org/10.1017/S095457940800059X

Peterson, M.J., Benca, R.M., 2006. Sleep in mood disorders. Psychiatr. Clin. 29, 1009–1032.

Qiu, M.-H., Vetrivelan, R., Fuller, P.M., Lu, J., 2010. Basal ganglia control of sleep–wake behavior and cortical activation. Eur. J. Neurosci. 31, 499–507. https://doi.org/10.1111/j.1460-9568.2009.07062.x

Robinson, J.L., Laird, A.R., Glahn, D.C., Blangero, J., Sanghera, M.K., Pessoa, L., Fox, P.M., Uecker, A., Friehs, G., Young, K.A., Griffin, J.L., Lovallo, W.R., Fox, P.T., 2012. The functional connectivity of the human caudate: An application of meta-analytic connectivity modeling with behavioral filtering. NeuroImage 60, 117–129. https://doi.org/10.1016/j.neuroimage.2011.12.010

Smith, G.T., Cyders, M.A., 2016. Integrating affect and impulsivity: The role of positive and negative urgency in substance use risk. Drug Alcohol Depend., Emotion Regulation and Drug Abuse: Implications for Prevention and Treatment 163, S3–S12. https://doi.org/10.1016/j.drugalcdep.2015.08.038

Sowell, E.R., Thompson, P.M., Holmes, C.J., Jernigan, T.L., Toga, A.W., 1999. In vivo evidence for post-adolescent brain maturation in frontal and striatal regions. Nat. Neurosci. 2, 859–861.

Steinberg, L., 2010. A dual systems model of adolescent risk-taking. Dev. Psychobiol. 52, 216– 224. https://doi.org/10.1002/dev.20445

Tashjian, S.M., Goldenberg, D., Galván, A., 2017. Neural connectivity moderates the association between sleep and impulsivity in adolescents. Dev. Cogn. Neurosci. 27, 35– 44. https://doi.org/10.1016/j.dcn.2017.07.006

Telzer, E.H., Goldenberg, D., Fuligni, A.J., Lieberman, M.D., Gálvan, A., 2015. Sleep variability in adolescence is associated with altered brain development. Dev. Cogn. Neurosci. 14, 16–22. https://doi.org/10.1016/j.dcn.2015.05.007

Tervo-Clemmens, B., Quach, A., Calabro, F.J., Foran, W., Luna, B., 2020. Meta-analysis and review of functional neuroimaging differences underlying adolescent vulnerability to substance use. NeuroImage 209, 116476. https://doi.org/10.1016/j.neuroimage.2019.116476

van den Bos, W., Rodriguez, C.A., Schweitzer, J.B., McClure, S.M., 2015. Adolescent impatience decreases with increased frontostriatal connectivity. Proc. Natl. Acad. Sci. 112, E3765–E3774. https://doi.org/10.1073/pnas.1423095112

Wager, T.D., Waugh, C.E., Lindquist, M., Noll, D.C., Fredrickson, B.L., Taylor, S.F., 2009. Brain mediators of cardiovascular responses to social threat: part I: Reciprocal dorsal and ventral sub-regions of the medial prefrontal cortex and heart-rate reactivity. NeuroImage 47, 821–835. https://doi.org/10.1016/j.neuroimage.2009.05.043

Wang, G., Grone, B., Colas, D., Appelbaum, L., Mourrain, P., 2011. Synaptic plasticity in sleep: learning, homeostasis and disease. Trends Neurosci. 34, 452–463.

Whiteside, S.P., Lynam, D.R., 2001. The Five Factor Model and impulsivity: using a structural model of personality to understand impulsivity. Personal. Individ. Differ. 30, 669–689. https://doi.org/10.1016/S0191-8869(00)00064-7

Yang, F.N., Bronshteyn, M., Flowers, S.A., Dawson, M., Kumar, P., Rebeck, G.W., Turner, R.S., Moore, D.J., Ellis, R.J., Jiang, X., 2021. Low CD4 nadir exacerbates the impacts of APOE ε4 on functional connectivity and memory in adults with HIV. AIDS Publish Ahead of Print. https://doi.org/10.1097/QAD.0000000000002840

Yang, F.N., Liu, T.T., Wang, Z., 2022a. Functional connectome mediates the association between sleep disturbance and mental health in preadolescence: A longitudinal mediation study. Hum. Brain Mapp. 43, 2041–2050. https://doi.org/10.1002/hbm.25772

Yang, F.N., Xie, W., Wang, Z., 2022b. Effects of sleep duration on neurocognitive development in early adolescents in the USA: a propensity score matched, longitudinal, observational study. Lancet Child Adolesc. Health. https://doi.org/10.1016/S2352-4642(22)00188-2

Yu, M., Linn, K.A., Cook, P.A., Phillips, M.L., McInnis, M., Fava, M., Trivedi, M.H., Weissman, M.M., Shinohara, R.T., Sheline, Y.I., 2018. Statistical harmonization corrects site effects in functional connectivity measurements from multi-site fMRI data. Hum. Brain Mapp. 39, 4213–4227. https://doi.org/10.1002/hbm.24241

Zhuang, X., Gross, C., Santarelli, L., Compan, V., Trillat, A.-C., Hen, R., 1999. Altered Emotional States in Knockout Mice Lacking 5-HT1A or 5-HT1B Receptors. Neuropsychopharmacology 21, 52–60. https://doi.org/10.1016/S0893-133X(99)00047-0

